# The Impact of a Night Travel Ban Policy on Road Traffic Accidents: Interrupted Time Series evidence in Zambia

**DOI:** 10.1101/2021.02.16.21251202

**Authors:** M Kantu Moonga, Peter Hangoma

## Abstract

The burden of road traffic accidents has been increasing globally with Injuries and deaths from road traffic accidents accounting for a significant share of the global disease burden. This is evident especially in low and middle income countries (LMICs), were these injuries and deaths account for a reasonable share of the disease burden. In Zambia, for example, road traffic accidents are the third leading cause of death after HIV/AIDS and Malaria and more than half of these accidents happen at night. To reverse the growing incidence of road crushes, the government of Zambia put a ban on night travel for public service vehicles in November, 2016. While other countries such as Kenya have implemented similar bans, there is no evidence on the extent to which such a ban may reduce accidents.

**Objective:** to examine the effect of the night travel ban on road traffic accidents in Zambia after the implementation of the night travel ban. The study set out to established whether there was a change in the number of accidents.

**Design:** the study design was a single group interrupted time series analysis. Administrative data on road traffic accidents in Zambia for the period 1964 to 2018 was used.

**Setting:** this research was a national study, therefore it encompassed national statistics on road accidents of the entire country of Zambia.

**Main outcome measure:** The total counts of road traffic accidents in Zambia recorded during the study period 1964 to 2018.

**Results:** it was found that the night travel ban reduced the number of road crushes by 1,211 within one year of implementing the intervention. (p value 0.001, CI −1878.079 to −543.130).

**Conclusion:** the night travel ban may be an effective way of reducing the burden of road traffic injuries in Zambia and other LMICs.

Section 1: What is already known on this topic

- In Zambia, there is a high number of road traffic accidents that occur during the night compared to the day time (fisa et al, 2019, ikabungo 2015 and patel 1979).
- Interventions have been put in place to reduce the number of accidents but there is no evidence of their effectiveness.
- In light of the lack of evidence of such interventions, the study was carried out to provide evidence to policy makers so that their decisions can be evidence based.

Section 2: what the study adds

- Our study suggests that the policy proved to be effective in reducing the total number of accidents in Zambia.
- This study provides evidence of the effectiveness of an intervention which in turn provides policy makers with grounds on which to maintain such a policy.
- The study triggers further research on other effects of banning night time travel

**Copyright/licence for publication:** The Corresponding Author has the right to grant on behalf of all authors and does grant on behalf of all authors, a worldwide licence to the Publishers and its licensees in perpetuity, in all forms, formats and media (whether known now or created in the future), to i) publish, reproduce, distribute, display and store the Contribution, ii) translate the Contribution into other languages, create adaptations, reprints, include within collections and create summaries, extracts and/or, abstracts of the Contribution, iii) create any other derivative work(s) based on the Contribution, iv) to exploit all subsidiary rights in the Contribution, v) the inclusion of electronic links from the Contribution to third party material where- ever it may be located; and, vi) licence any third party to do any or all of the above.”

## INTRODUCTION

### Background

The burden of Road Traffic Accidents (RTAs) has been growing globally, with the rate of mortality increasing by 46% since 1990 (Lozano et al., 2012). RTAs are responsible for nearly 20 to 50 million injuries each day and killing about 1.35 million people each year [World Health Organisation (WHO) 2018, 2020]. In 2020 it is expected that the number of deaths from accidents will reach 2 million (Mathers and Loncar, 2005, Wegman 2017). Road traffic injuries (RTIs) are ranked as the 8^th^ leading cause of death for all age groups worldwide and it is the leading cause of death for people between 5 – 29 years. More people die now from RTAs and RTIs than from HIV/AIDS [Centre for Disease control (CDC) 2019]. RTIs account for 83 million Disability Adjusted Life Years (DALYs) globally placing it among the 5 leading causes of DALYs (WHO GHO, 2020). Perhaps more concern is the fact that more than 90% of the DALYs lost worldwide due to RTIs occurred in low and middle income countries (LMICs) and yet they only account for 54% of registered global vehicles (WHO, 2015a). This underscores the importance of assessing what policies are needed to reduce RTAs in LMICs. Apart from the large health burden, RTAs also lead to large economic costs in terms of lost productivity and survivors of trauma go on to bear large health costs. A study of 21 countries on the global cost of RTIs estimated that they cost US$518 billion (Bachani et al., 2017). RTIs have a huge impact on health and development as they cost governments approximately 3% of Gross Domestic Product (WHO, 2015).

A number of countries have put in place measures to reduce the burden of RTAs. Measures include stricter rules on speed limits, laws on drunk-driving, seat belt use and improving road infrastructure (WHO 2001). Improving road infrastructure and related aspects such as lighting has shown to be important for road safety at night (Plainis et al., 2006). Others have banned travel in certain places or at certain times for vehicles or users deemed to be high risk. Countries such Kenya and Zambia have instituted night travel bans. Night travel has been associated with increased risk of RTAs in England (Horne, 1995), France (Nabi et al., 2006), Ethiopia (Teferi et al., 2014), China (Zhao, 2010), and Zambia (Ikabongo, 2015), among others. There is a rich literature that has looked at the impact of, or how, different policy measures are associated with RTAs. Staton et al. (2016) found that the most common intervention evaluated was legislation and it had the best outcomes when combined with strong enforcement initiatives or as part of a multifaceted approach. Speed control was also crucial to crash and injury prevention and road improvement interventions in LMICs settings which should carefully consider how the impact of improvements will affect speed and traffic flow. The study noted that further road traffic injury prevention interventions should be performed in LMICs with patient-centred outcomes in order to guide injury prevention in these complex settings. Grundy et al., (2009) examined the impact of an enforcement of 20 miles per hour (mph) traffic speed zones on RTAs and found a 41.9% reduction in road casualties. Navoa et al., (2011) evaluated the impact of penalty points system on road traffic injuries in Spain. The penalty point is a system where drivers start with a 12 to 10 point licence and points are gradually removed if certain traffic violations are committed. The study found a significant risk reduction in the post-intervention period with a greater risk reduction among men, moped riders and on urban roads. Porchia et al, 2014 found that visibility aids such as road lighting have the potential to reduce RTAs. There is limited or no evidence on the impact of night travel bans. The closest of evidence is an ex-ante study of the proposed policy to ban express buses from travelling in the early hour in Malaysia (Mohamed et al., 2011). The study concluded that a night travel ban may actually result in undesirable effects. Driving hours would be reduced and working hours would be affected because travellers will rush out of work to travel back home before the ban hours. There would be need for alternative transport for those who travel at night or long journeys and, resting places would be required for drivers. However, the findings were not based on the actual impact of the policy because it was carried out before the pronouncement. Their assessment was a likely impact if such a policy was implemented. In addition, the study used qualitative methods and this does not provide us with a picture of how the number of RTAs change. Other studies suggest that night travel bans may be helpful because night travel carries increased risk. Taylor et al., (2006) notes that a complex link exists between stress, fatigue, sleep, health status and drivers’ accident risk. The hormone melatonin which is associated with sleep and is usually suppressed during the day time but released at night (Masters et al., 2014), could be another reason explaining the high number of night time accidents. When released, melatonin causes sedation and is the reason why people feel the need to sleep at night. This can be cause for accidents when caught up on the road. We contribute in filling the gap in the literature examining the impact of policies to reduce RTAs in general by examining the impact of a night travel ban using a unique opportunity provided by a night travel ban of public transport in Zambia.

### Context

Over the past 50 years, Zambia has witnessed a transformation in the transport system as a result of both political and economic changes. This brought about liberalisation of the transport sector and has led to an increase in the number of vehicles such as minibuses, mid heavy buses and taxis, against limited growth of road infrastructure (Mubanga, 2014). Zambia has made major investments in the road sector. In 2012 the government embarked on a huge program of improving the road infrastructure (Road Development Agency, 2014). Generally, the public service vehicles such as buses remain a principle mode of transport for people and account for most of the traffic related fatalities. Fisa et al (2019) found that in Zambia, there is a reduced incidence of dying if one is using a private vehicle compared to a public vehicle. There are approximately 2,000 annual fatalities from RTAs recorded in Zambia (GRZ, 2015). The capital city, Lusaka, accounts for more than half the total number of nationwide RTAs (RTSA 2017). According to both historical and current studies, most accidents in Zambia occur during the night. In one such study, Patel (1979) found that 40% of the accidents occurred between 18.00 to 24.00 hours, 39% occurred between 24:00 to 12:00 hours and 21% occurred between 12:00 to 18:00 hours. A 2013 study showed the same trend with 53% of accidents happening at night and a death rate of 52.27% (Ikabongo, 2015). Fisa et al (2019) also concluded that driving in the early hours of the day particularly between 1am to 6am, had an increased incidence of death form traffic crashes. The financial costs that come with RTAs and RTIs in Zambia approaches 3% of its Gross Domestic Product (Schartz, 2008). RTAs cost the country at least K5 billion per year (Mwamba, 2018). Studies have also shown that injuries have been associated with reduced earned income and lower consumption. Injuries increase consumption in the health sector leading to an increase in health expenditure (Hangoma et al., 2017). This is a constraint on resources which could be channelled to other developmental programs and poverty alleviation. To curb the increasing number of accidents at night, in November 2016, Zambia instituted Statutory Instrument No 76 of 2016, the road traffic (public service vehicles) (restriction on night driving) banning night travel for all public service vehicles between 9pm and 6am (GRZ 2016).

## METHODS

Our study adopted a natural experimental design, particularly a single group Interrupted Time Series Analysis (ITSA) as a method of analysis. This is a time series analysis where the series is divided or interrupted by an intervention into two time periods, the pre-intervention period and the post-intervention period. The two-time period are compared to come up with a conclusion on the effects of the intervention. We analysed secondary data on the number of RTAs recorded annually, between 1964 and 2018 from the Zambia police road traffic unit and the road transport and safety agency. The study population included all the RTAs reported and recorded throughout the country in the study period. We also conducted robustness checks to ensure that the data met the prescribed assumptions of ITSA.

### Data and Variables

The main explanatory variable was the intervention: the introduction of the night travel ban in 2016. We compared the pre-intervention period (1964 – 2016 prior to the policy pronouncement in November) to the post intervention period (2017 to 2018). The data used from the year 1964 to 2018 gave us 55 time points. Our main outcome was the yearly counts of RTAs in Zambia. These included all accidents reported to have occurred in a year and recorded by the Zambia police service. 55 observations were available for both the pre and post intervention period. Data on RTAs segregated between night and day time accidents in the stated timeframe would have been ideal to create an intervention and control group, but the statistics were not available in this form. We also included other covariates such as motor vehicle population and human population in an alternative specification. It couldn’t be included in the main specification because this data was only available from 2005 to 2018 (14 time points). The variable motor vehicle population is the total number of vehicles registered with the road transport and safety agency in a year. Human population included the total number of people recorded in Zambia each year and this data was collected for the study period 1964 to 2018. The design was aimed at showing whether there was an interruption in the trend after the night travel ban. We noted that history is the most common threat to validity with the possibility that some other event could have caused the observed effect in time series. In this regard, we also included dummies in the years Zambia liberalised the transport sector to account for possible structural breaks. We also carried out robustness checks.

### Empirical Model

To investigate the effect of the night travel ban on the number of RTAs, we began with an interrupted time series model of the form:

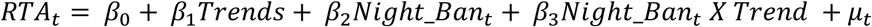

Where *RTA*_*t*_ is the number of accidents in year t. Trend is the time trends equal to 1 in the first year, 2 in the second year and so on counting the number of years from 1964 to 2018. It controls for trends. *Night_Ban*_*t*_ is a dummy variable equal to 1 in the period after the night travel ban (post-intervention period) and 0 in the pre-intervention period. It captures the intervention. *Night_Ban*_*t*_ *X Trend* is the interaction between the Night Travel Ban and RTAs over time. *μ*_*t*_ is the error term capturing all other factors that may affect RTAs. ß_0_represents the starting level of RTAs in 1964. ß_1_ is the trajectory of RTAs from the starting level in 1964 until the time that the Night Travel Ban was effected in 2016. The parameter ß_2_ represents the change in the level of RTAs that occurred in the period immediately following the introduction of the Night Travel Ban compared with the counterfactual. The counterfactual in this case is the predicted slope of progression of RTAs through 2017-18, had the intervention not been put in place in 2016. ß_3_ describes the difference between the pre-intervention slope and the post-intervention slope of RTAs. The parameters of interest in our study are ß_2_ and ß_3_. Figure i below shows model (1) graphically:

**Fig i:**
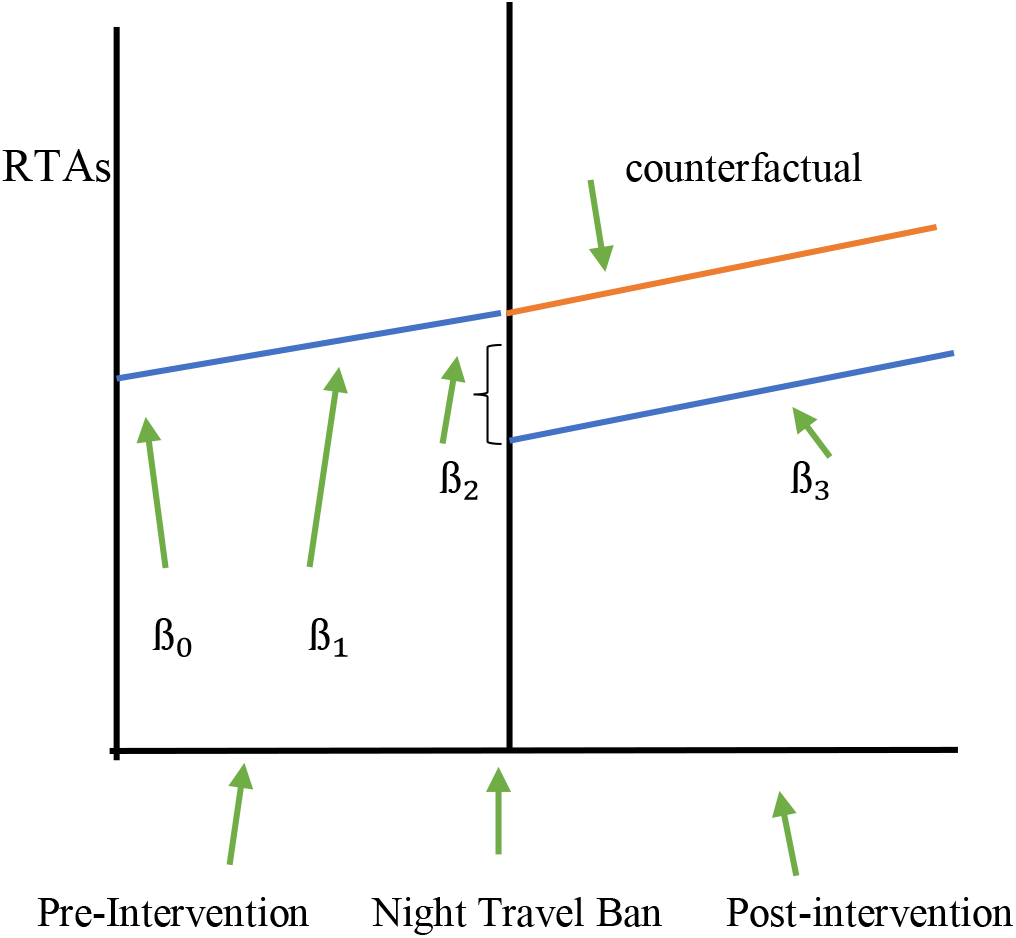
Single Group Interrupted Time Series of the Impact of RTAs in Zambia. Where ß_0_ is the starting point of RTAs in1964, ß_1_ is the change in RTAs over time up until the ban in 2016, ß_2_ measures the change in the number of RTAs after the ban is implemented while ß_3_ measures the change of RTAs over time in the period that the ban is in place.

The model was further modified to include a vector *Z*_*t*_ that included the number of motor vehicles recorded, human population (number of people) and structural breaks.

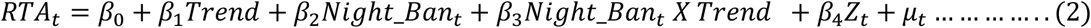

Structural breaks were defined as year dummies for most of the periods in the 1990s and early 2000s to account for liberalization of the public transport sectors and other reforms.

Finally, we assessed the model to check for stationarity, normality, homoskedasticity and autocorrelation. This was carried out to make sure that important assumptions about the consistency of these observations were met and that the estimates were the Best Linear Unbiased Estimators (BLUE). This was done to ensure that the study design was robust (Linden A,2016).

## RESULTS

### Descriptive statistics and trends in RTAs

The number of RTAs, motor vehicle population and human population are described through 1964 to 2018 (Table 1). During the 55 year study period, the average annual number of RTAs that have occurred is 13,745 with a minimum of 3,680 and maximum of 33, 672 (standard deviation 8,111). These RTAs were further disaggregated into sub-periods to create further understanding on the trend, and these sub-periods were randomly created. The average number of RTAs between 1964 and 1973 is 8,120; between 1974 and 1991 is 8,650; between 1992 and 2000 is 12,314 and; between 2000 and 2018 is 22, 651. These results show a steady increase of RTAs over the 55 year period under study. Motor vehicle population had 14 observations and the average number of motor vehicles recorded in the country in the last 14 years is 455,677.8 with the maximum of 788,168 motor vehicles. 55 observations (1964-2018) were included for human population and they showed an average population of 8,824,621 with a maximum of 17,600,000.

**Table 1:**
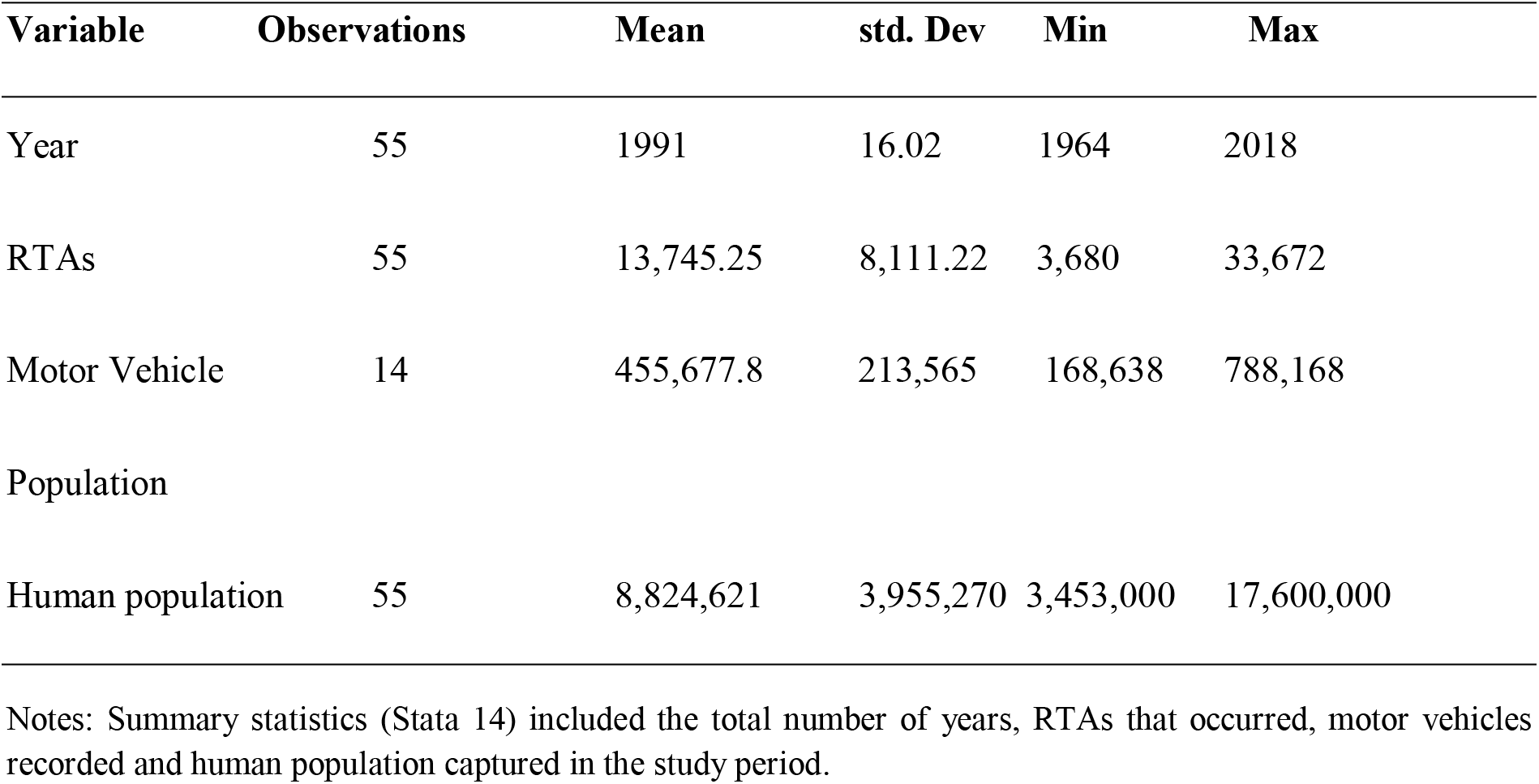
Summary Statistics.

| Variable | Observations | Mean | std. Dev | Min | Max |
| --- | --- | --- | --- | --- | --- |
| Year | 55 | 1991 | 16.02 | 1964 | 2018 |
| RTAs | 55 | 13,745.25 | 8,111.22 | 3,680 | 33,672 |
| Motor Vehicle | 14 | 455,677.8 | 213,565 | 168,638 | 788,168 |
| Population |  |  |  |  |  |
| Human population | 55 | 8,824,621 | 3,955,270 | 3,453,000 | 17,600,000 |

Notes: Summary statistics (Stata 14) included the total number of years, RTAs that occurred, motor vehicles recorded and human population captured in the study period.

Figure 2 is a graphical depiction of the analysis of RTAs through the study period and it shows an upward trend of RTAs characterised with some fluctuations. The interrupted time series model includes a trend to account for this. The period of 1995 to 2003 showed a more defined fluctuating pattern as the trend followed a sharp upward and downward trend. The fluctuation corresponds to a period of transport liberalization and hence the need to include structural breaks in the model. The dotted vertical line is the intervention depicting the year 2016 when the Night Travel Ban was effected. Visually, the graph showed a reduction in the number of RTAs after the intervention was put in place as evidenced in Table 2. The predicted number of RTAs is plotted against the actual number of RTAs recorded.

**Table 2:**
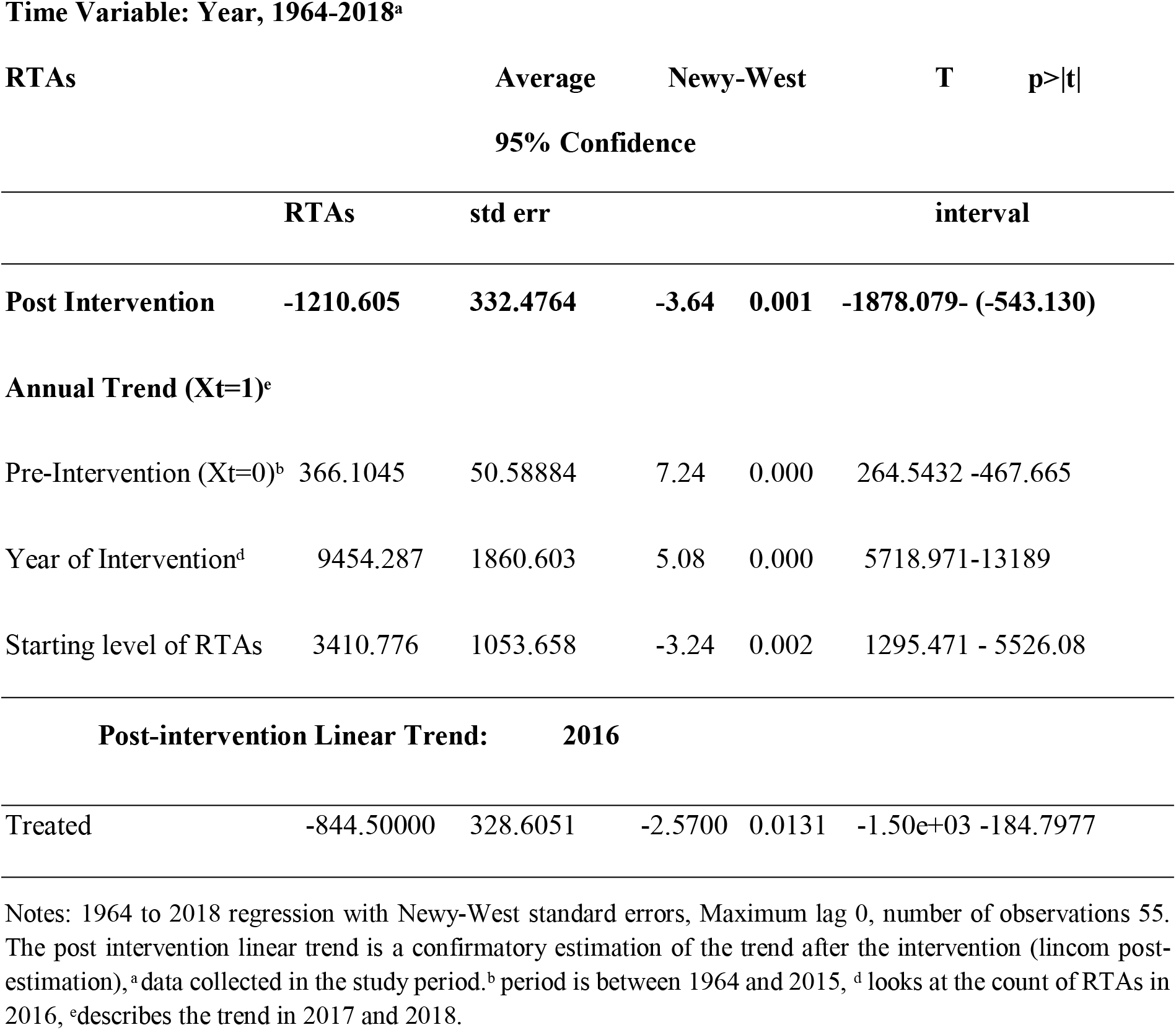
Effect of the night travel ban on road accidents in Zambia.

**Table 3:** Summary of ITSA with motor vehicle and human population.

| **Controls** | **Pre- intervention** | **intervention** | **post- intervention** |
| --- | --- | --- | --- |
|  | **(1964-2015)** | **(2016)** | **(2017-2018)** |
| Non (1964-2018) | increase in RTAs | increase in RTAs | reduction in RTAs |
| Population (1964-2018) | reduction in RTAs | **** | reduction in RTAs |
| Vehicles (2005-2018) | increase in RTAs | reduction in RTAs | reduction in RTAs |

\*\*\*\* Results were not significant

**Table 4:** Number of RTAs per 100,000 of the population.

| **Year** | **number of RTAs/ 100,000 population** |
| --- | --- |
| 2015 | 2,172 |
| 2016 | 2,022 |
| 2017 | 1,774 |
| 2018 | 1,742 |

**Figure 2:**
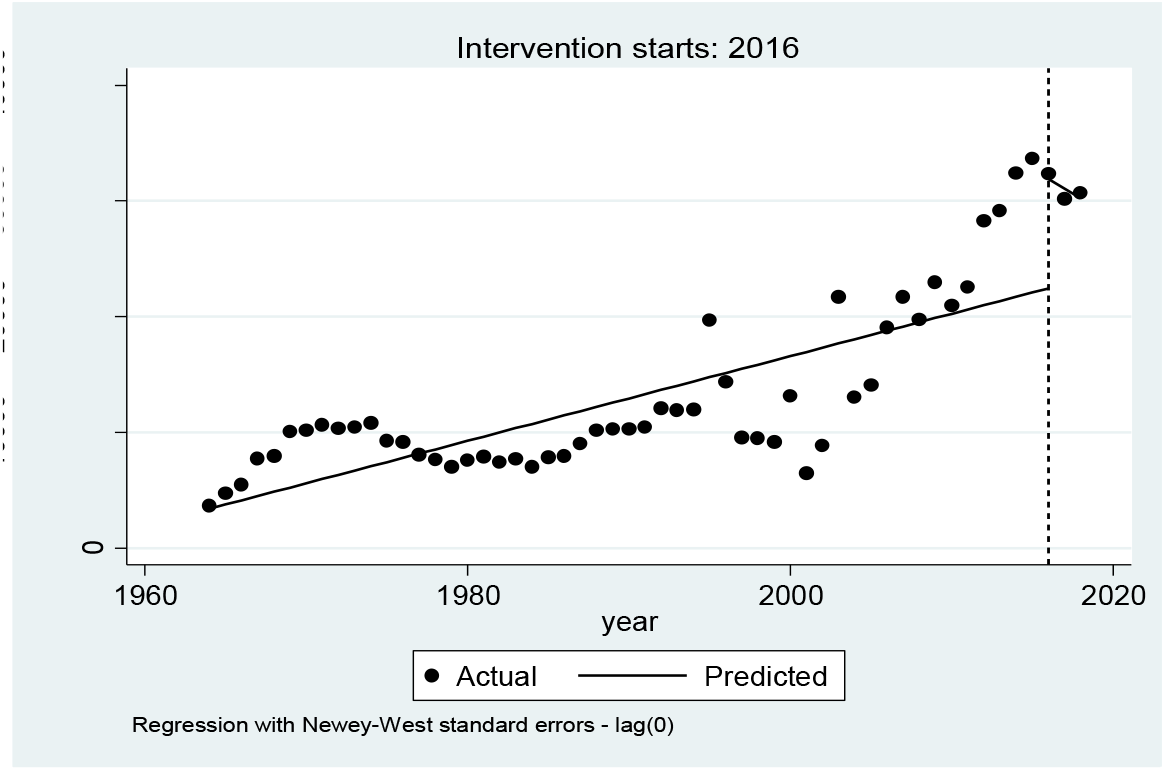
RTAs and intervention (2016)

**Figure iii:**
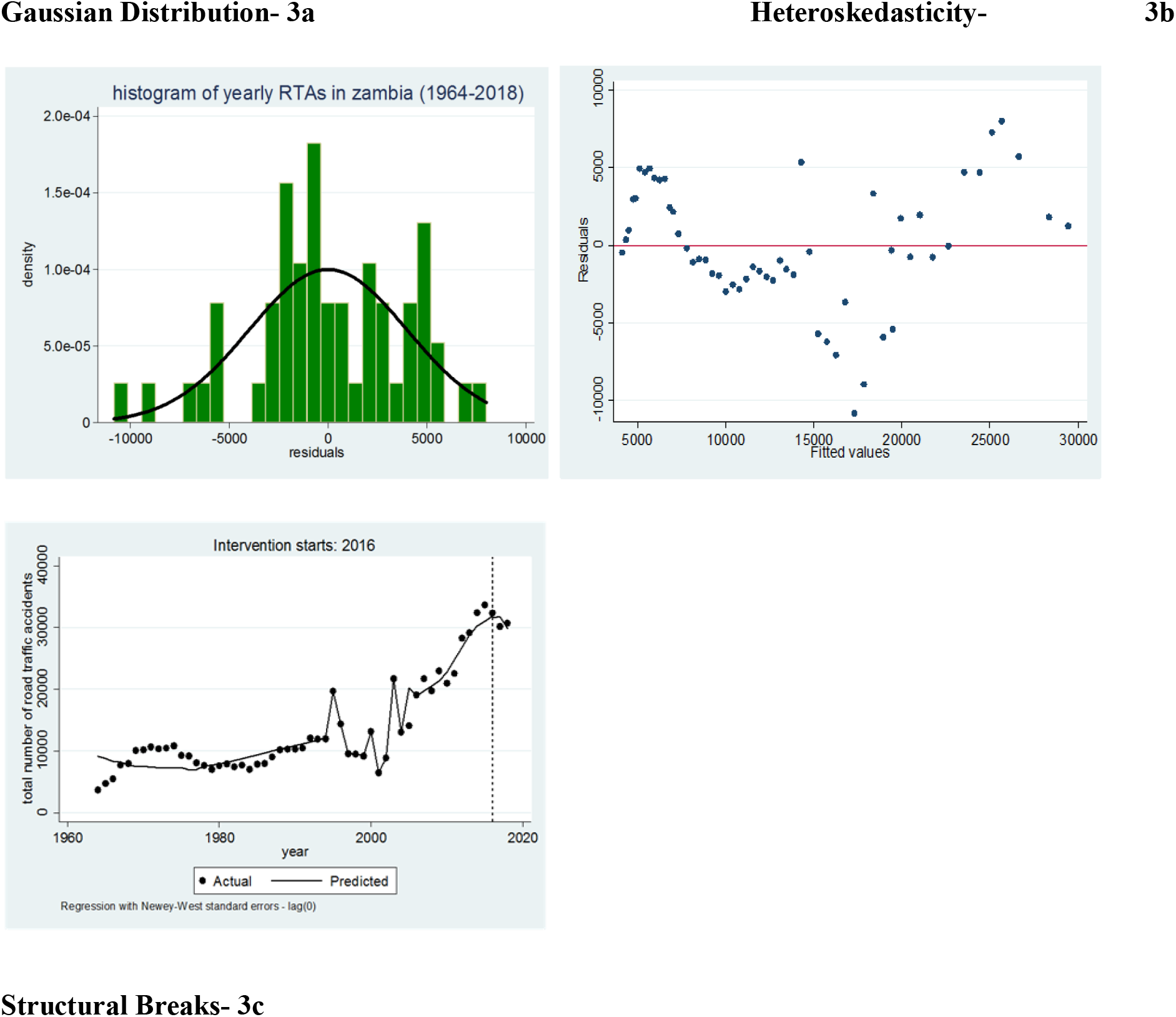
Diagnostics.

### The Effect of the Night Travel Ban on RTAs in Zambia

Results show that the night travel ban was associated with a decrease in RTAs by 1.211(Table 2, Row 1). The decrease was highly significant at below the convectional 1% level. Looking at other parameters of the model, the number of RTAs increased significantly every year prior to the intervention by an average of 366. In the first year of the intervention (2016), the analysis showed an increase of RTAs by approximately 9,454 (p value 0.0001, CI = 5718.971, 13189).

When human population was put into consideration, the analysis showed an increase in RTAs before and in the year of the intervention (1964-2016), then a decrease in RTAs in the post intervention period (2017-2018). Further the analysis showed a reduction in RTAs in the intervention period (2016-2018) when we controlled for motor vehicle population.

When the RTAs were analysed per 100, 000 of the population (Table iv), the analysis showed a decreasing trend in the pre-intervention period (2015), intervention period (2016) and post-intervention period (2017-2018). In a time series analysis, a post estimation is carried out to confirm the trend of an analysis. The Lincom Post-estimation was employed and it confirmed the reduction of RTAs after the intervention.

### Robustness Checks

We also checked if the data met the assumptions of time series data. The robustness tests for stationarity (pre-estimation) and normality, homoscedasticity, autocorrelation, structural breaks (post-estimation) were carried out. The test for stationarity (Dickey Fuller test) was positive. In the post estimation, the residuals showed a normal distribution (skewness and kurtosis tests). The residuals had characteristics of Heteroscedasticity (8b) and we corrected this by carrying out the analysis using the robust standard error. We also identified autocorrelation (Durbin-Watson statistics, Breush-Godfrey test). This was corrected by transforming the Durbin-Watson statistic. We also checked for structural breaks. The analysis was finally carried out with all these corrections and the results of the analysis still showed a significant reduction in RTAs. We concluded that the estimators were best linear unbiased estimators.

## DISCUSSION

The finding shows a reduction in the number of RTAs in Zambia after implementation of the intervention. We attribute the reduction in RTAs to the Night Travel Ban since it was an intervention that was put in place in that time period. Other interventions may have also contributed to this reduction but this analysis could not take them into account individually. The analysis however shows that RTAs are actually preventable if the correct interventions such as a ban on night travel are put on place. The ban on night travel received a different review in a study carried out in Malaysia prior to the policy pronouncement. Mohammed et al (2011) in his impact assessment analysis concluded that a ban on night time driving would bring about negative effects in the agenda against RTAs. Other studies have shown the effectiveness of improving conditions for people driving at night in order to reduce the number of RTAs. Plainis et al., (2006) also established that corrective measures on night time driving such as increasing lighting reduced RTAs. In Zambia the problem of infrastructure particularly street lighting in rural areas where villagers walk in the main road in the late evening, has contributed to night time accidents involving pedestrians, cyclists and motorists (Schatz, 2008). Night time driving has shown to have more RTAs despite the reduction in the total number of vehicles or the number of miles driven at night compared to day time. Night time driving has certain characteristics. It has fatigued drivers and other factors came into play such as reduced visibility, poor judgement and delayed reaction time. A ban on night time driving would however be more effective in reducing RTAs if a more holistic approach would be considered. Drivers of PSVs should be encouraged to rest and therefore, they and their passengers should have designated resting places if they are caught up in transit during the time of the ban. There is also concern on the economic impact this ban has on the travellers who could be travelling at night to meet productive deadlines. Alternative means of transport should be considered for passengers who need to travel at night such as rail and air transport.

The method of analysis itself (ITSA), shows the ability to demonstrate the change in the trend at a defined point in time resulting from an intervention is a tested method. This method is also cost effective and efficient in evaluating national based interventions which might be too expensive to evaluate. The study provided a lesson in both the pre and post intervention periods and such methodological study designs have the capacity to impact on risk reduction policy interventions.

## CONCLUSION

We found that the banning of public service vehicles from moving at night lead to a reduction in the total number of road traffic accidents recorded in Zambia. The number of RTAs took an upward trend before the intervention was put in place and later showed a downward trend in the period of the intervention. Evidence of an intervention that is effective has been presented in this study. The analysis had a couple of limitations. Secondary data faces challenges of potential lack of accuracy in recording. There could also have been non-reported or recorded RTAs, especially in rural areas. The study could not take into account other road safety measures that may have contributed to the reduction in RTAs in this period. There was also no specified data on the type of motor vehicles involved in these accidents since the treatment was instituted on the PSVs. The statistics were not available on a quarterly or monthly basis. This would have shown how the trend differs from month to month or quarter to quarter and given a picture of factors that influence such differences.

## Data Availability

data is available on request

## ABREVIATIONS

RTAs: Road traffic accidents
PSVs: Public Service Vehicles
ITSA: Interrupted Time Series Analysis
DALYs: Disability Adjusted Life Years Lost
WHO: World Health Organisation
RTIs: Road Traffic Injuries
ZRST: Zambia Road Safety Trust

## ACKNOWLEDGEMENTS

The study benefited from data collected from the Zambia police service and the road transport and safety agency. We thank the faculty at the school of public health at the University of Zambia for their continued advice.

## DATA SHARING

Data collected on this study can be shared on request.

